# ^18^FDG Positron Emission Tomography Mining for Metabolic Imaging Biomarkers of Radiation-induced Xerostomia in Patients with Oropharyngeal Cancer

**DOI:** 10.1101/2020.05.17.20104737

**Authors:** Hesham Elhalawani, Carlos E. Cardenas, Stefania Volpe, Souptik Barua, Sonja Stieb, Calvin Rock, Timothy Lin, Pei Yang, Haijun Wu, Jhankruti Zaveri, Baher Elgohari, Lamiaa E. Abdallah, Amit Jethanandani, Abdallah S.R. Mohamed, Laurence E. Court, Katherine A. Hutcheson, G. Brandon Gunn, David I. Rosenthal, Steven J. Frank, Adam S. Garden, Arvind Rao, Clifton D. Fuller

## Abstract

**Purpose:** Head and neck cancers (HNC) radiotherapy (RT) is associated with inevitable injury to parotid glands and subsequent xerostomia. We investigated the utility of standardized uptake values (SUV) derived from routinely performed 18-fluorodeoxygluocose positron-emission tomography (^18^FDG-PET) to develop metabolic imaging biomarkers (MIBs) of RT-related parotid injury.

**Methods:** Data for oropharyngeal cancer (OPC) patients treated with RT at our institution between 2005-2015 with available planning computed tomography (CT), dose grid, pre- & first post-RT ^18^FDG-PET-CT scans, and physician-reported xerostomia assessment at 3-6 months post-RT (Xero 3-6ms) per CTCAE, was retrieved, following an IRB approval. A CT-CT deformable image co-registration followed by voxel-by-voxel resampling of pre & post-RT ^18^FDG activity and dose grid were performed. Ipsilateral (Ipsi) and contralateral (contra) parotid glands were sub-segmented based on the received dose in 5 Gy increments, i.e. 0-5 Gy, 5-10 Gy sub-volumes, etc. Median and dose-weighted SUV were extracted from whole parotid volumes and sub-volumes on pre- & post-RT PET scans, using in-house code that runs on MATLAB. Wilcoxon signed-rank and Kruskal-Wallis tests were used to test differences pre- and post-RT.

**Results:** 432 parotid glands, belonging to 108 OPC patients treated with RT, were sub-segmented & analyzed. Xero 3-6ms was reported as: non-severe (78.7%) and severe (21.3%). SUV- median values were significantly reduced post-RT, irrespective of laterality (p=0.02). A similar pattern was observed in parotid sub-volumes, especially ipsi parotid gland sub-volumes receiving doses 10-50 Gy (p<0.05). A Kruskal-Wallis test showed a significantly higher mean planned RT dose in the contra parotid in the patients with more severe Xero 3-6mo (p= 0.03). Multiple logistic regression showed a combined clinical-dosimetric-metabolic imaging model could predict the severity of Xero 3-6mo; AUC=0.78 (95%CI:0.66-0.85;p<0.0001)

**Conclusion:** We sought to quantify pre- and post-RT ^18^FDG-PET metrics of parotid glands in patients with OPC. Temporal dynamics of PET-derived metrics can potentially serve as MIBs of RT-related xerostomia in concert with clinical and dosimetric variables.

## Introduction

Head and neck cancer (HNC) radiotherapy (RT) is associated with inevitable injury to the parotid glands due to the radiation beam pathways, with subsequent xerostomia. Xerostomia is also the most often reported radiation-induced side effect in these patients, with 50% exhibiting acute Grade (G) 2/3 and 32% late G2/3 xerostomia. [1] This can further contribute to other radiation-induced symptoms like dysphagia, speech problems, and taste alteration, in addition to secondary problems like dental caries. [2-4] Although there is a potential for recovery even years after RT, xerostomia remains a major detrimental factor to patients’ quality of life after HNC RT. [5] With the broad use of positron emission tomography (PET) as an imaging modality to assess treatment response in HNC, [6, 7] the question arises if PET can also be used to quantify changes in organs at risk after radiotherapy and to assess the degree of xerostomia. [8-10]

In this study we therefore investigated the utility of standardized uptake values (SUV) derived from routinely performed fluorodeoxyglucose PET (^18^FDG-PET) scans for staging and response evaluation to develop metabolic imaging biomarkers (MIBs) of RT-related parotid injury. The specific aims of our study can be summarized as follows:

1. Outline an analytical workflow for radiotherapy-associated normal tissues toxicities assessment studies incorporating PET imaging
2. Investigate the dose-response relationship between RT dose and longitudinal alterations of quantitative PET SUV metrics (Δ changes) in the parotid gland
3. Assess the utility of Δ SUV changes to model the severity of subacute RT-induced parotid injury and subsequent xerostomia at 3-6 months.

## Methods

### Study Population

Following an approval from an institutional review board (IRB) at the University of Texas M.D. Anderson Cancer Center, data for biopsy proven oropharyngeal cancer (OPC) patients treated between 2005 and 2015 who underwent radiation therapy as a single or multimodality definitive therapy were considered for the current investigation (n=150). This investigation and relevant methodology were performed in compliance with the Health Insurance Portability and Accountability Act (HIPAA) as a retrospective study where the need for informed consent was waived [11]. Electronic medical records were scanned for various demographic, disease, and treatment characteristics in the absence of any prior head and neck re-irradiation. (**Table 1**) The aspects of our institutional multidisciplinary approach for managing oropharyngeal cancer patients -including RT planning-were previously reported in detail. [12, 13] Eligibility criteria required individuals to have received pre-treatment and post-RT ^18^FDG-PET-CT scans, and have retrievable planning CT and dose grid, as well as Xerostomia assessment at 3-6-months following RT course start (Xero 3-6mo). Common Terminology Criteria for Adverse Events (CTCAE v5.0) were applied to grade Xerostomia using a Likert scale that ranged between 0 (no) and 3 (severe). [14]

### PET-CT acquisition protocol and eligibility criteria

Each patient underwent two ^18^FDG-PET/CT scans, the first within 4 weeks prior to starting therapy and the second within 3-6 months following RT course initiation (median interval of around 135 days). FDG-PET/CT images were uniformly acquired and analyzed with a single scanner (Discovery ST-8; GE Medical Systems, Milwaukee, WI), as previously described. [15]

### Image registration and parotid glands segmentation

After CT-CT deformable image co-registration using commercial software (Velocity AI). Then resampling of pre & post-RT ^18^FDG-PET scans and dose grid was done so that these scans matched the pixel and slice spacing of the treatment planning CT (**Figure 1**). Ipsilateral (ipsi) and contralateral (contra) parotid glands were sub-segmented into smaller sub-volumes based on the dose they received using 5Gy increments, *i.e*. 0-5Gy, 5-10Gy sub-volumes, *etc*. These sub-volumes were automatically defined using in-house software (MATLAB, MathWorks, Natick, MA) which extracted each glands’ contoured masks and overlaid these structures with each patient’s isodose lines to extract the desired sub-volumes (Figure 2). Using the treatment plans’ dose map and isodose lines, we define the sub-volumes by identifying the union between parotid structure and the voxels within the desired isodose lines. These dose sub-volumes were defined on the pre & post-RT CT scans (from PET/CT) after deformably registering these scans to the treatment planning CT scan.

**Fig. 1.**
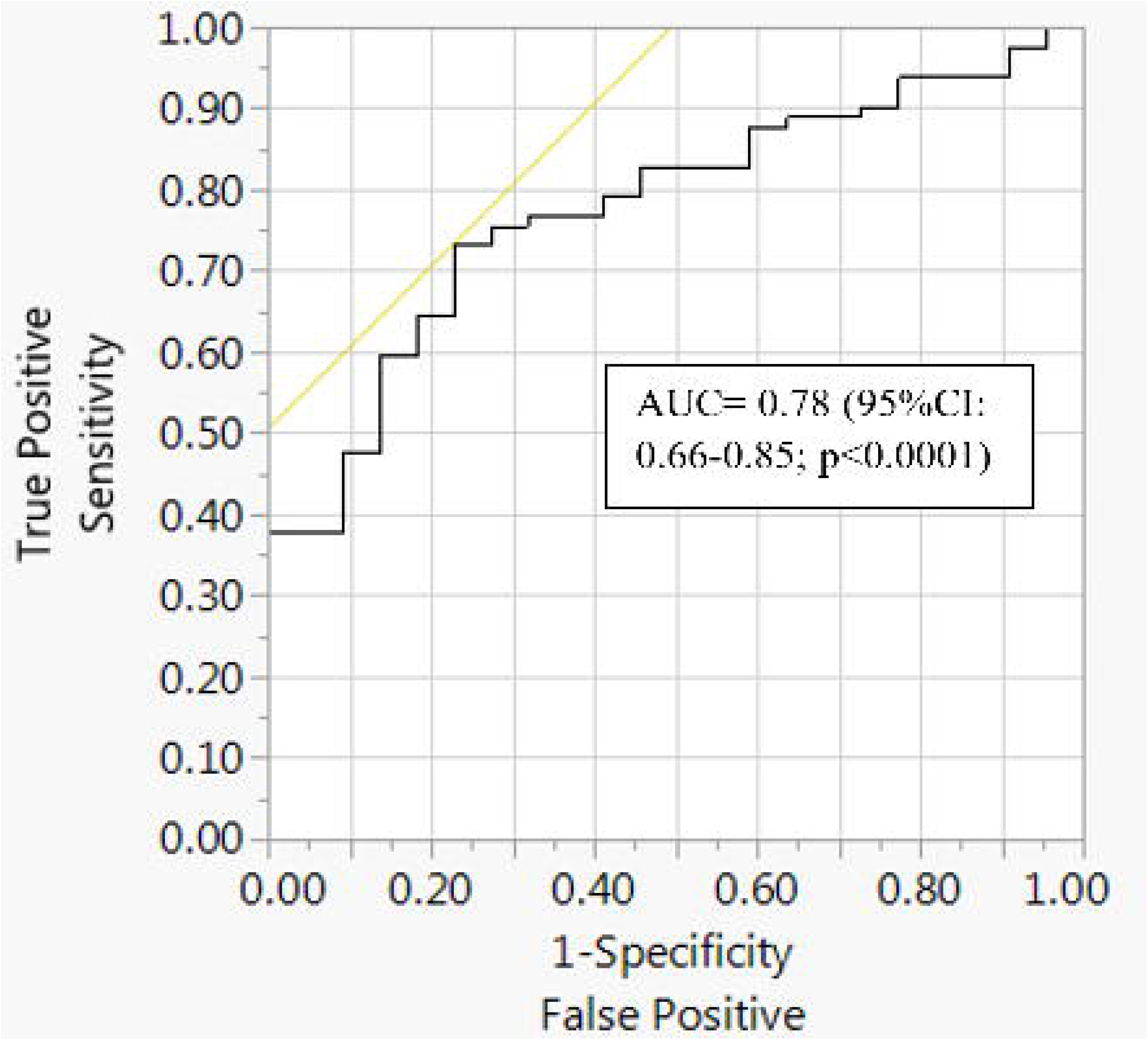
Parotid dose sub-volume generation. Using the treatment plans’ dose map and isodose lines, we define the sub-volumes by identifying the union between parotid structure (white) and the voxels within the desired isodose lines. For example, for the parotid sub-volume (yellow) receiving doses between 5 and 10Gy, between 10 and 15Gy, and between 65 and 70Gy are shown with their respective isodose lines for each panel

**Fig. 2.**
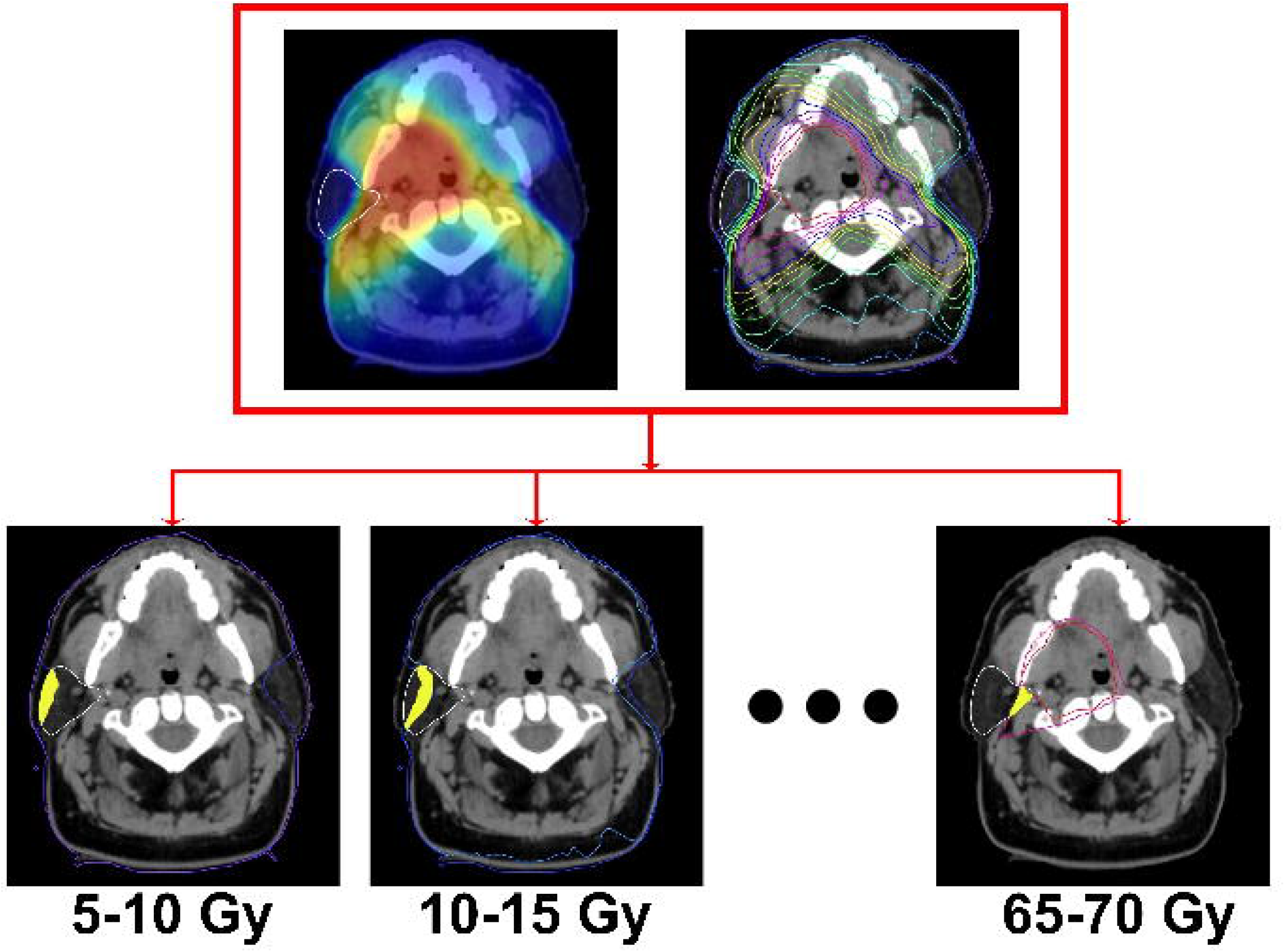
Illustration of dose-weighted SUV computation. For each voxel in the co-registered image space, the SUV map and dose map matrices are multiplied using the Hadamard product [46] resulting in the dose-weighted SUV (DwSUV) maps

### ^18^FDG-PET metabolic features extraction

Absolute and dose-weighted median SUV were extracted from these whole parotid glands and sub-volumes on pre- & post-RT PET scans, using in-house software. Dose-weighted SUV is defined by the voxel-wise multiplication of the registered PET scan’s SUV map and radiotherapy planned dose map (**Equation 1**),

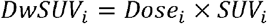

where *i* represents each voxel in the image space. An illustration of the dose-weighted SUV is shown in **Figure 2**.

### Statistical analysis

Differences between median values of SUV-median pre-and post-RT for the whole glands volumes and sub-volumes were tested using Wilcoxon signed-rank test. We then applied Spearman’s Rho rank correlation analysis to compute the correlation coefficient and corresponding p-values to find if percent changes in values on a whole or sub-volume level are significantly correlated with mean accumulated dose at the relevant volume of interest. Single then multiple logistic regressions were used to model the correlation between clinical, dosimetric, and SUV variables (e.g. age, induction chemotherapy, and baseline xerostomia) and Xero 3-6mo. A p value of <0.05 was considered significant. 95% CI of ROC AUC was obtained by 10,000 bootstrapping.

## Results

### Patients

432 parotid glands, belonging to 108 patients with OPC treated with RT, were delineated, sub-segmented & analyzed. Assessments of xerostomia at 3-6 months following initiation of radiation treatment were available for 105 patients and were reported as: non-severe, i.e. CTCAE grades 0, 1, and 2 (78.7%) and severe, i.e. CTCAE grade 3 (21.3%). Various demographics, disease, and treatment characteristics are reported in **Table 1**.

**Table 1.** Patients, disease and treatment characteristics

| Characteristics | N (%) |
| --- | --- |
| **Sex** |  |
| Male | 96 (88.89%) |
| Female | 12 (11.11%) |
| Age at diagnosis, years: median (interquartile range 'IQR')* | 59.3 (53.4-64) |
| **Tumor laterality** |  |
| Right | 50 (46.3%) |
| Left | 58 (53.7%) |
| **Oropharynx subsites** |  |
| Base of tongue | 57 (52.8%) |
| Tonsil | 41 (38%) |
| Others | 10 (9.2%) |
| **T category** |  |
| T0 | 3 (2.8%) |
| T1 | 32 (29.6%) |
| T2 | 47 (43.5%) |
| T3 | 16 (14.8%) |
| T4 | 9 (8.4%) |
| Tx | 1 (0.9) |
| **N category (ICON-S)** |  |
| N0 | 1 (0.9%) |
| N1 | 72 (66.7%) |
| **N2** | 33 (30.6%) |
| **N3** | 2 (1.8%) |
| **Therapeutic combination** |  |
| **Radiation alone** | 12 (11.1%) |
| **Induction chemotherapy (IC) followed by concurrent chemoradiation** | 28 (25.9%) |
| **IC followed by radiation alone** | 17 (15.7%) |
| **CC** | 51 (47.3%) |
| **Radiation dose (median; IQR) [Gy]\*** | 70 (66-70) |
| **Radiation fractions (median; IQR)** | 33 (30-33) |
| **Baseline xerostomia** |  |
| **Yes** | 23 (21.3%) |
| **No** | 82 (75.9%) |
| **Not reported** | 3 (2.8%) |
| **Xerostomia at 3-6 months (CTCAE v5.0)\*** |  |
| **0** | 1 (0.9%) |
| **1** | 45 (41.7%) |
| **2** | 36 (33.3%) |
| **3** | 23 (21.3%) |
| **Not reported** | 3 (2.8%) |
| **Mean parotid dose (Gy; standard deviation)** |  |
| **Ipsilateral** | 35.4 (13.1) |
| **Contralateral** | 19.7 (10.4) |

\*IQR: inter-quartile range; Gy: Gray; CTCAE v5.0: 5<sup>th</sup> version of common terminology criteria for adverse events.

### Parotids dosimetry and SUV metrics analytics

Ipsi and contra parotid glands received mean doses ‘Gy’ (Std) of 35.4 (13.2) & 20 (10.4), respectively. SUV-median tends to decrease significantly after RT (p= 0.02) for ipsi and contra parotid glands, where 62% and 57% of the parotid glands, respectively demonstrated a decline in SUV-median post-therapy, respectively. **(Supplementary Table S1)**

We also observed a similar pattern of variation in SUV metrics across time on a parotid sub-volume level. Post-RT SUV-median values of parotid sub-volumes which received doses ‘5-50 Gy’ decreased significantly as compared to their pre-RT counterparts, on combining both sides. This mainly applied to ipsi parotid gland sub-volumes receiving doses ‘10-50 Gy’ (p<0.05). **(Supplementary Table S2)**

A Spearman rank correlation test showed that overall; percent change in SUV-median was not correlated to mean accumulated dose at parotid glands. Similarly, negligible correlation between the sub-volumes median SUV changes and mean or discrete dose levels was found. This suggests that higher doses do not imply a higher magnitude of percent changes in SUV metrics, and vice-versa.

### Modeling of Xerostomia using clinical, dosimetric, and metabolic imaging features

Three patients with no recorded xerostomia assessment at 3-6 months were excluded (n=3). For the remaining 105 patients, Xero 3-6mo was graded per CTCAE v5.0 as: 0 (1%), 1 (42.9%), 2 (34.3%), and 3 (21.9%). For the purpose of our analysis, we recategorized Xero 3-6mo into: non-severe (CTCAE G<3; 78.1%) and severe (CTCAE G3; 21.9%). A Kruskal-Wallis test showed a significantly higher mean planned RT dose in the contra parotid in the subgroup who suffered from more severe Xero 3-6mo (p= 0.03). Patients with more severe Xero 3-6mo were also shown to have higher mean planned ipsi parotid glands doses (p= 0.1), and lower Post-RT parotid SUV-median both for ipsi (p=0.04) and contra (p=0.02) parotid glands. Nevertheless, neither absolute nor dose-weighted percent changes in SUV-median correlated to Xero 3-6mo severity. Older age was also associated with more sever Xero 3-6mo (p= 0.01). Distribution of other clinical variables like therapeutic combination, AJCC stage (8^th^ ed.), or presence of pre-treatment xerostomia didn’t significantly differ between patients with severe and non-severe Xero 3-6mo. (**Figure 3**)

**Fig. 3.**
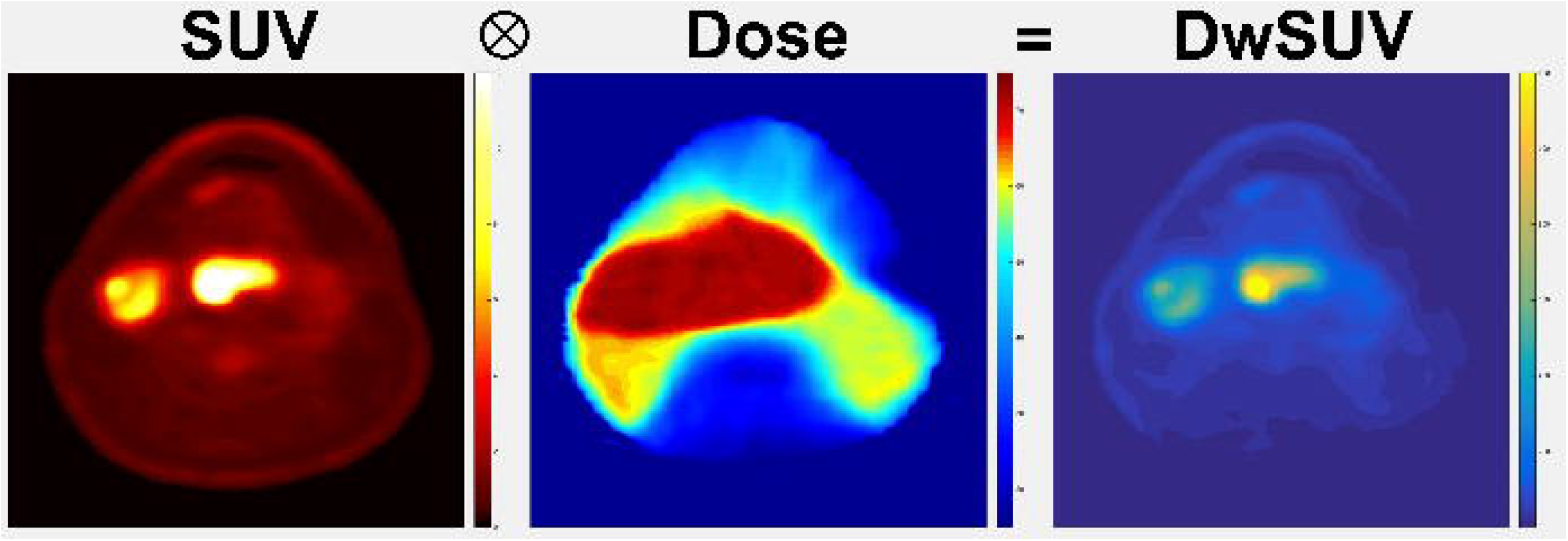
Correlation between severity of post-radiotherapy xerostomia and (A) Age; (B) Mean contralateral parotid gland dose (Gy); (C) Post-radiotherapy contralateral parotid gland SUV-median; and (D) Post-radiotherapy ipsilateral parotid gland SUV-median

A simple logistic regression (SLR) of individual clinical and dosimetric co-variates -known to affect salivary function and subsequent response to RT injury- in addition to extracted SUV metrics was performed. Older age (p=0.04), higher mean planned RT dose at contra parotid glands (p=0.001), and post-RT ipsi (p=0.03) and contra (p=0.1) parotid SUV median were significantly correlated to Xero 3-6mo. Though non-significant, higher mean ipsi parotid gland dose (p=0.07) and more advanced cancer stage (p=0.1) predicted more severe Xero 3-6mo.

We then performed a multiple logistic regression analysis including only statistically significant variables on SLR. Overall, this combined clinical-dosimetric-metabolic imaging model could adequately predict the severity of Xero 3-6mo as evidenced by an AUC of 0.78 (95%CI: 0.66-0.85; p<0.0001), on plotting an ROC. (**Figure 4**) Contra parotid glands post-RT SUV-median and mean planned RT dose stood out as the most statistically significant variables (p=0.03 for both) followed by age at diagnosis (p=0.06). Notably, mean ipsi parotid gland dose was detrimental to the predictive performance of this model as evidence by a 4-point decline in BIC upon its omission. (**Supplementary Table S3**).

**Fig. 4.**
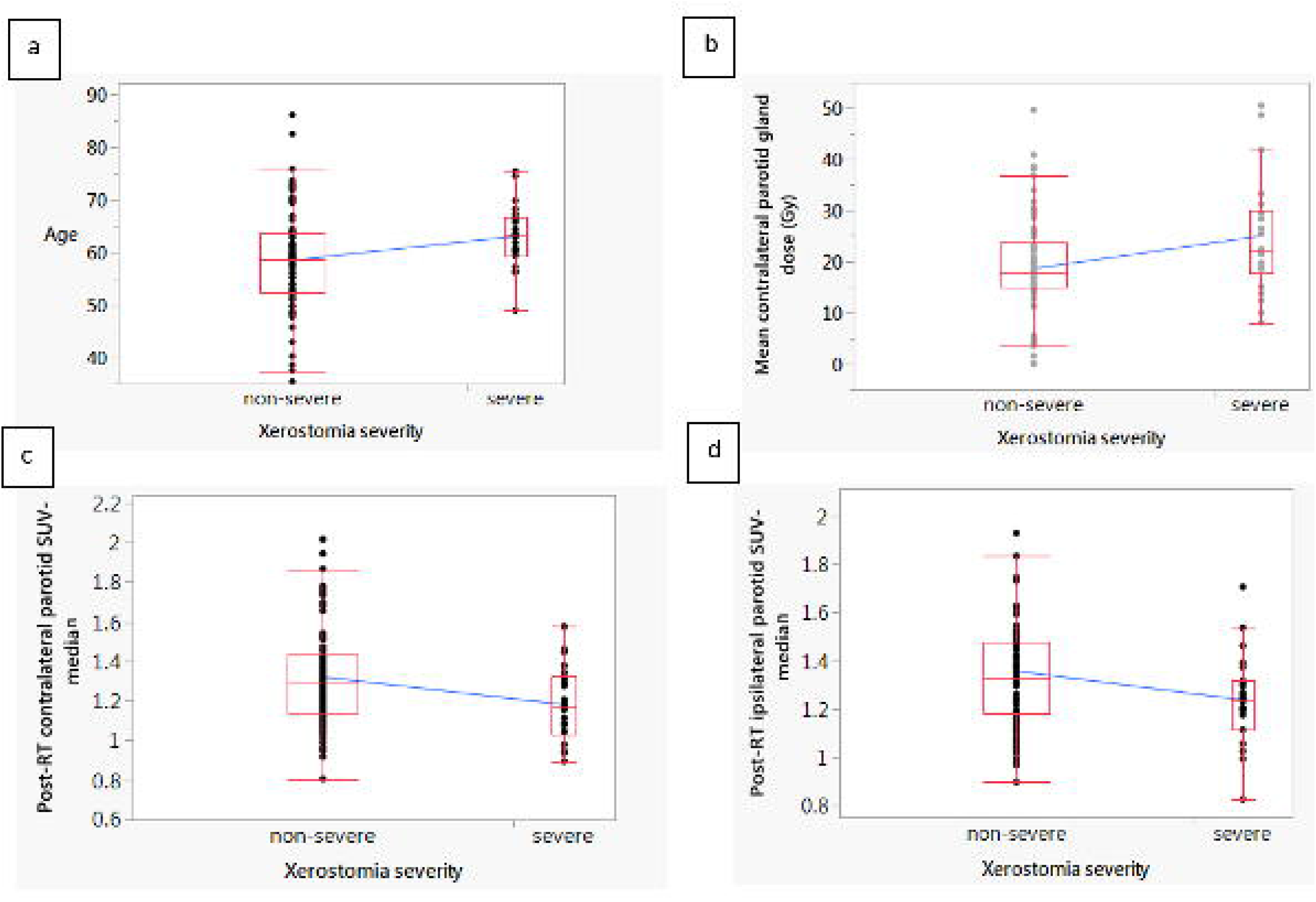
Receiver Operator Characteristic Curve (ROC) displaying the model performance represented by area under the curve (AUC)

## Discussion

In this study, we show that the median SUV of both, ipsilateral and contralateral parotid glands, decrease significantly after radiotherapy compared to baseline values. This was valid for nearly all dose levels, showing that even after very small radiation doses a change in SUV can still be detected. One possible explanation of the decrease in median SUV is the reduction in acinar cells [16-19] with simultaneous increase of intercellular water or fat, which can be visualized in magnetic resonance imaging (MRI) by an increase in T2 [20, 21] or ADC [22], and a rising fat fraction in DIXON MRI. [20] The direct negative correlation between ADC and FDG uptake [23] has been confirmed in several studies using a simultaneous PET/MR in pre-treatment HNC [24], lung cancer [25], lymphoma [26], and liver metastases [27] or post-RT as in rectal cancer [28] and retroperitoneal fibrosis. [29]

Only five other studies have previously investigated the change in SUV of the parotid gland from pre- to post-radiotherapy (**Table 2**). Direct comparison between these studies and our results is challenging due to large differences in methodology and analysis. In our study, we only used the median SUV of the parotid glands for analysis. This was considered due to the fact that the SUV of the parotid gland can be largely affected by PET-positive lymph nodes in direct vicinity to the gland. Tumoral spread to these level II lymph nodes is specifically the case in locally advanced oral cavity, naso- and oropharyngeal cancers, which are among the most common head and neck tumors. [30] Not only the partial volume effect due to the voxel size matters, but also -and maybe more importantly- the halo around the FDG active tumor tissue, which can account for several millimeters, depending on the SUV window level. [31, 32] This can have significant effect on the maximum SUV, and therefore also affects the mean SUV. To avoid at least the partial volume effect of surrounding tissue on the PET results, Buus et al. cropped their parotid gland structure by 5mm. This was a sophisticated approach but resulted in some parotid gland volumes as low as 4ml. [33] Unfortunately, subsequent PET image analysis of the parotid gland, published by other study groups, did not consider this confounding factor, although including lymph node positive oropharyngeal tumors in most cases (**Table 2**). Hence, the reported pre-treatment mean and maximum SUV values were most probably overestimated, whereas -due to tumor shrinkage- the post-treatment values would, if at all, only be slightly affected. Both will result in a falsely overestimated decrease in SUV mean and maximum after therapy, which will represent an effect of radio- and/or chemotherapy on the tumor rather than on the parotid gland.

**Table 2.** Overview of studies analyzing PET as biomarker of radiation-induced injury of the parotid glands. The comparison with our study is mainly hampered due to the missing correction for PET halo of FDG positive level II lymph nodes in most other studies (OP: oropharynx; L: larynx; HP: hypopharynx; NP: nasopharynx; OC: oral cavity; FDG: fluorodeoxyglucose; PET: positron emission tomography; RT: radiotherapy; IMRT: intensity modulated RT; VMAT: volumetric modulated arc therapy; w: weeks; m: months; BL: baseline; SUV: standardized uptake value; sign.:significantly; LNs: lymph nodes; HNC: head and neck cancers; NA: not applicable)

| Author, year | N | Tumor entity | Node-positive disease | RT technique | PET tracer | Time point PET | Major findings<br>Positive aspects/major limitations |
| --- | --- | --- | --- | --- | --- | --- | --- |
| Elhalawani, 2019 (this study) | 108 | OP | 99% | IMRT | 18F-FDG | BL<br>3-6m post-RT start (median 135 days) | Median SUV of ipsi- and contralateral parotid gland sign. decreased after RT; lower median SUV values post-RT (ipsi or post?), higher mean dose to the contralateral parotid gland and age sign. associated with severe xerostomia post-RT<br>Pros: Patient number; OPC only; Correction for partial volume effects and SUV halo of level II LNs |
| Van Dijk, 2018 [34] | 161 | HNC (OP, L, HP, NP, OC) | 56% | IMRT (90%) / VMAT (10%) | 18F-FDG | BL | Xerostomia at BL, higher mean dose to the parotid gland and lower median and mean SUV of the parotid gland at BL were sign. associated with higher risk for xerostomia at 12 months (unclear if ipsi-, contralateral or both parotid glands)<br>Pros: Patient number; prospective assessment of xerostomia at 12m<br>Cons: No full correction for partial volume effects and SUV halo of level II LNs; only BL PET imaging; dose received by the parotid glands unclear; no comparison between BL xerostomia and SUV |
| Cannon, 2012 [9] | 98 / 14 | HNC (OP, L, HP) | 84%/93% | IMRT | 18F-FDG | BL<br>7-9w post-RT | Mean fractional parotid SUV (= SUV post-tx / pre-tx; unclear if ipsi-, contralateral or both parotid glands) of 0.96 (range 0.62 – 1.85); stimulated saliva decreased post-RT to 41% relative to BL; neg. correlation between xerostomia grade and fractional SUV<br>Pros: Subgroup of patients with prospectively assessed sialometry and xerostomia-specific questionnaire<br>Cons: Unclear which SUV value has been used (i.e. median, mean, max); no correction for partial volume effects and SUV halo of level II LNs; xerostomia assessment in only 8 patients |
| Roach, 2012 [46] | 49 | HNC (OP, L, CUP, NP) | NA | IMRT | 18F-FDG | BL<br>6-151w post-RT (mean 22w) | Mean SUVmean decreased by 5% with every 10 Gy increase in mean parotid gland dose; decrease in mean SUVmax by 8% with mean dose to the parotid gland of $\leq 20$ Gy, and 46% with doses $> 50$ Gy<br>Cons: No correction for partial volume effects and SUV halo of level II LNs; no xerostomia assessment; wide range of PET FU |
| Buus, 2006 [33] | 12 | HNC (L, OP, CUP, HP, OC) | 50% | Ipsi-wedged pair (3) OP (6), 3D (2), IMRT (1) | 11C-methionine | 8-54m post-RT (median 21m) | Net metabolic clearance of 11C-methionine neg. correlated with RT dose to the parotid gland; TD50 of 30 Gy (individual variation from 7 – 50Gy)<br>Pros: Correction for partial volume effects; correlation of net metabolic clearance of 11C-methionine and RT dose in parotid gland sub-volumes (5 Gy intervals)<br>Cons: PET tracer not used in clinical routine; only 1 patient with IMRT and without hypoxic cell sensitizer; wide range of PET FU; low patient number |
| Buus, 2004 [47] | 8 | HNC (OP, CUP) | 75% | Ipsi(5), IMRT (1), NA(2) | 11C-methionine | BL (n=2)<br>6-29m post-RT (median 22m; n=6) | Higher 11C-radioactivity concentration in salivary glands (parotid and submandibular glands) receiving lower RT doses; net metabolic clearance of 11C-methionine neg. correlated with RT dose and pos. correlated to salivary gland function<br>Pros: Saliva collection; dynamic PET scan<br>Cons: PET tracer not used in clinical routine; no correction for partial volume effects and SUV halo of level II LN; only 1 patient with IMRT and without hypoxic cell sensitizer; different patients at baseline and post-RT; wide range of PET FU; low patient number |
| Rege, 1993 [48] | 11 | HNC (OP, Sinus, OC, NP, L) | 40% | NA; Additional brachytherapy (2) | 18F-FDG | BL During RT 1-10w post-RT (usually 6w) | No sign. difference in FDG uptake between salivary glands „in field“ or „out of field“, and no sign. change with RT<br>Pros: Analysis of parotid, submandibular and sublingual glands; assessment also during RT; FDG uptake normalized on healthy cerebral tissue<br>Cons: Parotid gland contour on only one slice pre-treatment, which has been copied and rotated to fit post-treatment scan; no correction for partial volume effects and SUV halo of level II LNs; dose received by the “in field” parotid glands unclear; 2 patients with additional brachytherapy; early post-RT PET scan; not the same patients for all time points; no correlation to xerostomia; low patient number |

Hence, a reasonable comparison of our study results could only be performed with the above-mentioned study of Buus et al. [33] However, this study used ^11^C-methionine as PET tracer and only one of their 12 patients received IMRT without a hypoxic cell sensitizer not to mention they reported the net clearance of the tracer rather than SUV values. The main finding of their study is the dose-dependency of the net clearance described as a sigmoid pattern. In our study however, although nearly 10 times more patients were studied, we couldn’t see a dose dependency of the PET changes. One possible explanation could be the different median time interval between radiotherapy and PET scan which was 21 months in the study by Buus et al. and 3-6 months in our study. Probably, high doses to the parotid gland will result in prolonged damage, whereas low dose areas have the potential to still recover or recover faster from the RT-induced damage over time. To validate this, a longitudinal PET study needs to be conducted or more data from other studies need to be available. From MRI studies, we know that recovery of the parotid glands, measured as percent changes in volume or ADC values, most probably occurs after 6-8 months from end of RT. [22]

On stratifying patients according to severity of xerostomia at 3-6 months post-RT, we found significantly lower median SUV values post-RT in the ipsi and contra parotid glands, higher mean contralateral parotid gland doses and a more elderly population in the patient subgroup that suffered from more severe symptoms. Of note, neither the pre-treatment median SUV, nor the percentage change in median SUV significantly correlated with xerostomia at 3-6 months, indicating that the post-treatment SUV is the most reliable PET image biomarker for assessment of xerostomia at that time point. Of note, none of the pre-RT SUV metrics were significantly associated with baseline xerostomia. However, van Dijk et al. could show a negative correlation of the baseline SUV values with xerostomia at twelve months post-RT. [34] Noteworthy, they haven’t analyzed the correlation between baseline SUV values and baseline xerostomia, and did not perform a second measurement during or post-RT.

Moreover, in our study a model combining post-RT median SUV of contralateral parotid gland with age and mean dose to contralateral parotid gland could correlate with the severity of post-RT xerostomia with demonstrable accuracy (AUC = 0.78). This shows that PET-derived imaging biomarkers can potentially play a role in evaluation of RT-induced normal tissue injury as previously demonstrated by our group using longitudinal CT and MRI studies. [35-40]

Despite the strength of our study with the largest patient cohort having two PET assessments, before and after RT, inclusion of OPC patients only, compensation for partial volume effect and PET halo in the analysis and simultaneous physician-reported toxicity assessment - using a standardized tool (CTCAE v5.0) - with PET scan acquisition, our study has some limitations. First of all, the auto-contoured parotid glands have been manually adapted only in selected regions, where the program did not perform very well. However, with our approach described above, to analyze the median SUV only, this might have had affected the PET results only to a minor extent and only in the dose level sub-volume analysis, when small regions at the edges were analyzed. Second, we only analyzed SUV changes in dose level sub-volumes up to 50 Gy to exclude possible PET halo effects of nearby FDG positive lymph nodes. We have chosen 50 Gy as involved lymph node received at least 54 Gy and we assumed that the GTV to CTV/PTV margin and the dose falloff until 50 Gy will prevent the inclusion of any PET halo. Nevertheless, future analysis needs to be done to exactly assess the extent of such a PET halo and to find solutions for future analysis.

Another limitation is the fact, that although all PET scans have been performed within three to six months from the start of radiotherapy, they were not conducted at a standardized time point, potentially capturing parotid glands metabolic activity at varying recovery phases. However, contradicting results are available in the literature describing a decrease [41], increase [42] or stable values [43] for dry mouth between 2 and 6 months post-RT using the European Organisation for Research and Treatment of Cancer (EORTC) H&N35 questionnaire. Last, with the measurement after end of therapy, the ability to adapt the treatment according to changes in the FDG uptake is not possible anymore. Nevertheless, in our study we took the diagnostic post-treatment images for analysis to avoid further radiation to the patient with an additional mid-treatment scan. However, in MRI, changes in the parotid gland can be detected already during radiotherapy and predict for late xerostomia [44, 45], so a prospective study with an additional mid-treatment PET scan seems to be justifiable.

In conclusion, temporal dynamics of PET-derived metrics manifest changes that can potentially serve as MIBs of RT-related parotid injury. Our results help generate hypothesis for the utility of integrating mid-RT PET-CT into adaptive RT trials for HNCs.

## Data Availability

We are currently working on anonymizing matched clinical and imaging datasets per HIPPA regulations prior to uploading the data to a public data repository

## Declarations

### Funding

There are no direct funding sources for this study to disclose

### Conflicts of interest/Competing interests

There are no conflicts of interest/competing interests to disclose

## Compliance with Ethical Standards

### Ethics approval

This retrospective chart review study involving human participants was in accordance with the ethical standards of the institutional and national research committee and with the 1964 Helsinki Declaration and its later amendments or comparable ethical standards. The Institutional Review Board (IRB) of the University of Texas MD Anderson Cancer Center approved this study. (RCR-03-0800).

### Consent to participate

The need for consent to participate was waived given the retrospective nature of the study

### Consent for publication

The need for consent to participate was waived given the retrospective nature of the study

### Availability of data and material (data transparency)

Not available yet

### Code availability (software application or custom code)

Not available yet

### Authors’ contributions

All authors whose names appear on the submission

1. made substantial contributions to the conception or design of the work; or the acquisition, analysis, or interpretation of data; or the creation of new software used in the work;
2. drafted the work or revised it critically for important intellectual content;
3. approved the version to be published; and
4. agree to be accountable for all aspects of the work in ensuring that questions related to the accuracy or integrity of any part of the work are appropriately investigated and resolved.

**Conceptualization:** Hesham Elhalawani, Carlos Cardenas, Stefania Volpe, Clifton Fuller; **Methodology:** Hesham Elhalawani, Carlos Cardenas, Stefania Volpe, Souptik Barua, Sonja Stieb, Calvin Rock, Timothy Lin, Pei Yang, Haijun Wu, Jhankruti Zaveri, Baher Elgohari, Lamiaa Abdallah, Amit Jethanandani, Abdallah Mohamed**; Formal analysis and investigation:** Hesham Elhalawani, Carlos Cardenas, Stefania Volpe, Souptik Barua, Abdallah Mohamed**; Writing - original draft preparation:** Hesham Elhalawani, Carlos Cardenas, Sonja Stieb**; Writing - review and editing:** Hesham Elhalawani, Carlos Cardenas, Stefania Volpe, Souptik Barua, Sonja Stieb, Calvin Rock, Timothy Lin, Pei Yang, Haijun Wu, Jhankruti Zaveri, Baher Elgohari, Lamiaa Abdallah, Amit Jethanandani, Abdallah Mohamed, Laurence Court, Katherine Hutcheson, G. Brandon Gunn, David Rosenthal, Steven Frank, Adam Garden, Arvind Rao, Clifton Fuller**; Supervision:** Laurence Court, Katherine Hutcheson, G. Brandon Gunn, Arvind Rao, Clifton Fuller.

## Acknowledgments

Dr. Elhalawani was supported in part by the philanthropic donations from the Family of Paul W. Beach to Dr. G. Brandon Gunn, MD. Drs. Elhalawani and Fuller receive funding and project-relevant salary support from NIH/NCI Head and Neck Specialized Programs of Research Excellence (SPORE) Developmental Research Program Award (P50 CA097007-10). This research is supported by the Andrew Sabin Family Foundation; Dr. Fuller is a Sabin Family Foundation Fellow. Drs. Fuller and Mohamed receive funding and project-relevant salary support from the National Institutes of Health (NIH), including the National Institute for Dental and Craniofacial Research Award (1R01DE025248-01/R56DE025248-01); National Cancer Institute (NCI) Early Phase Clinical Trials in Imaging and Image-Guided Interventions Program(1R01CA218148-01); and National Science Foundation (NSF), Division of Mathematical Sciences; NIH Big Data to Knowledge (BD2K) Program of the National Cancer Institute Early Stage Development of Technologies in Biomedical Computing, Informatics, and Big Data Science Award (1R01CA214825-01). Dr. Fuller receive funding and project-relevant salary support from NIH/NCI Cancer Center Support Grant (CCSG) Pilot Research Program Award from the UT MD Anderson CCSG Radiation Oncology and Cancer Imaging Program (P30CA016672) and National Institute of Biomedical Imaging and Bioengineering (NIBIB) Research Education Program (R25EB025787). Dr. Fuller has received direct industry grant support and travel funding from Elekta AB. Dr. Fuller is supported via a National Science Foundation (NSF), Division of Mathematical Sciences, Joint NIH/NSF Initiative on Quantitative Approaches to Biomedical Big Data (QuBBD) Grant (NSF 1557679). Dr. Barua and Dr. Rao were supported by CCSG Bioinformatics Shared Resource P30 CA016672, CPRIT RP170719, CPRIT RP150578, NCI 1R37CA214955-01A1, a gift from Agilent technologies, and a Research Scholar Grant from the American Cancer Society (RSG-16-005-01). Dr. Elgohari is funded through joint supervision program by the Egyptian Ministry of Cultural and Higher Education. None of these industrial partners’ equipment was directly used or experimented within the present work.

## Supplementary tables

**Supplementary Table S1.** Comparison of median values (interquartile range: IQR) of whole parotid glands SUV-median pre- and post-radiotherapy

| Parotid gland laterality | Pre-RT median values | Post-RT median values | p-value* | Delta (%) | % with declining SUVmedian |
| --- | --- | --- | --- | --- | --- |
| Ipsilateral parotid glands | 1.39 (1.19-1.61) | 1.31 (1.17-1.46) | 0.003 | -5.4% (-19.4% to 7.8%) | 67 (62%) |
| Contralateral parotid glands | 1.33 (1.13-1.57) | 1.28 (1.12-1.42) | 0.04 | -5.2% (-19.4% to 14.4%) | 61 (57%) |
| Bilateral parotid glands | 2.72 (2.33-3.13) | 2.55 (2.33-2.86) | 0.02 | NA* | NA |

\*NA: Not applicable

**Supplementary Table S2.** Comparison of median values of SUV-median derived from parotid sub-volumes pre- and post-radiotherapy

| Significant dose levels | Pre-RT median values | Post-RT median values | p-value* |
| --- | --- | --- | --- |
| **Bilateral parotid glands** |  |  |  |
| SUV-Median 0-5Gy | 1.15 (0.96-1.38) | 1.13 (0.96-1.30) | 0.18 |
| SUV-Median 5-10Gy | 1.32 (1.08-1.57) | 1.23 (1.03-1.40) | **0.01** |
| SUV-Median 10-15Gy | 1.34 (1.14-1.61) | 1.26 (1.12-1.44) | **0.004** |
| SUV-Median 15-20Gy | 1.39 (1.13-1.64) | 1.28 (1.13-1.43) | **0.002** |
| SUV-Median 20-25Gy | 1.39 (1.11-1.64) | 1.30 (1.12-1.47) | **0.004** |
| SUV-Median 25-30Gy | 1.38 (1.11-1.65) | 1.29 (1.13-1.47) | **0.002** |
| SUV-Median 30-35Gy | 1.41 (1.12-1.62) | 1.29 (1.13-1.46) | **0.0009** |
| SUV-Median 35-40Gy | 1.39 (1.13-1.58) | 1.29 (1.15-1.45) | **0.0013** |
| SUV-Median 40-45Gy | 1.38 (1.13-1.57) | 1.29 (1.15-1.44) | **0.003** |
| SUV-Median 45-50Gy | 1.36 (1.14-1.60) | 1.30 (1.16-1.45) | **0.02** |
| **Ipsilateral parotid glands** |  |  |  |
| SUV-Median 0-5Gy | 1.12 (1.02-1.31) | 1.09 (0.95-1.21) | 0.62 |
| SUV-Median 5-10Gy | 1.27 (1.06-1.52) | 1.17 (1.00-1.38) | 0.27 |
| SUV-Median 10-15Gy | 1.32 (1.17-1.58) | 1.25 (1.10-1.46) | **0.025** |
| SUV-Median 15-20Gy | 1.39 (1.13-1.59) | 1.27 (1.09-1.47) | **0.008** |
| SUV-Median 20-25Gy | 1.41 (1.15-1.64) | 1.28 (1.12-1.50) | **0.007** |
| SUV-Median 25-30Gy | 1.43 (1.14-1.67) | 1.31 (1.13-1.49) | **0.003** |
| SUV-Median 30-35Gy | 1.46 (1.17-1.64) | 1.32 (1.16-1.47) | **0.001** |
| SUV-Median 35-40Gy | 1.45 (1.21-1.65) | 1.33 (1.17-1.47) | **0.001** |
| SUV-Median 40-45Gy | 1.45 (1.21-1.63) | 1.34 (1.18-1.47) | **0.002** |
| SUV-Median 45-50Gy | 1.48 (1.19-1.66) | 1.35 (1.19-1.47) | **0.004** |
| **Contralateral parotid gland** |  |  |  |
| SUV-Median 0-5Gy | 1.16 (0.90-1.48) | 1.14 (0.96-1.33) | 0.38 |
| SUV-Median 5-10Gy | 1.36 (1.11-1.59) | 1.27 (1.07-1.43) | **0.012** |
| SUV-Median 10-15Gy | 1.41 (1.11-1.65) | 1.27 (1.14-1.44) | 0.07 |
| SUV-Median 15-20Gy | 1.38 (1.10-1.64) | 1.29 (1.15-1.42) | 0.10 |
| SUV-Median 20-25Gy | 1.36 (1.06-1.64) | 1.31 (1.13-1.43) | 0.16 |
| SUV-Median 25-30Gy | 1.36 (1.06-1.63) | 1.28 (1.12-1.44) | 0.16 |
| SUV-Median 30-35Gy | 1.33 (1.05-1.59) | 1.26 (1.11-1.43) | 0.16 |
| SUV-Median 35-40Gy | 1.34 (1.05-1.56) | 1.24 (1.12-1.42) | 0.23 |
| SUV-Median 40-45Gy | 1.33 (1.08-1.53) | 1.25 (1.13-1.41) | 0.26 |
| SUV-Median 45-50Gy | 1.29 (1.07-1.54) | 1.27 (1.14-1.43) | 0.79 |

\*p-values were generated using Wilcoxon-signed rank test

**Supplementary Table S3.** Variables effect summary represented by log worth and p-value

| Source | LogWorth | P-Value |
| --- | --- | --- |
| Post-radiotherapy contralateral parotid gland SUV-median | 1.580 | 0.02629 |
| Mean contralateral parotid gland dose (Gy) | 1.555 | 0.02787 |
| Age | 1.258 | 0.05515 |
| Mean ipsilateral parotid gland dose (Gy) | 0.379 | 0.41787 |

## Notes

### Competing Interest Statement

The authors have declared no competing interest.

### Clinical Trial

Not applicable. Retrospective study

